# Real-world uptake of COVID-19 vaccination among individuals expressing vaccine hesitancy: a registry-linkage study

**DOI:** 10.1101/2022.08.02.22278300

**Authors:** Kristin L. Andrejko, Jennifer F. Myers, Nozomi Fukui, Lauren Nelson, Rui Zhao, John Openshaw, James P. Watt, Seema Jain, Joseph A. Lewnard, Jake M. Pry, the California COVID-19 Case-Control Study Team

## Abstract

**Background:** Uptake of COVID-19 vaccination remains suboptimal in the United States and other settings. Though early reports indicated that a strong majority of people were interested in receiving the COVID-19 vaccine, the association between vaccine intention and uptake is not yet fully understood.

**Methods:** During 24 February-5 December 2021, we enrolled California residents receiving molecular tests for SARS-CoV-2 infection who had not yet received any COVID-19 vaccine doses. Unvaccinated participants provided information on their intentions to receive COVID-19 vaccination in a telephone-administered survey. We matched study participants with a state-wide immunization registry and fit a Cox proportional hazards model comparing time to vaccination among those unvaccinated at study enrollment by vaccination intention (willing, unsure, or unwilling).

**Findings:** Among 864 participants who were unvaccinated at the time of interview, 272 (31%) had documentation of receipt of COVID-19 vaccination later; including 194/423 (45.9%) who had initially reported being willing to receive vaccination, 41/185 (22.2%) who reported being unsure about vaccination, and 37/278 (13.3%) who reported unwillingness to receive vaccination. Adjusted hazard ratios (aHRs) for registry-confirmed COVID-19 vaccination were 0.49 (95% confidence interval: 0.32-0.76) and 0.21 (0.12-0.36) for participants expressing uncertainty and unwillingness to receive vaccination, respectively, as compared with participants who reported being willing to receive vaccination. Time to vaccination was shorter among participants from higher-income households (aHR 3.30 [2.02-5.39]) and who reported co-morbidities or immunocompromising conditions (aHR 1.54 [1.01-2.36]); time to vaccination was longer among participants who tested positive for SARS-CoV-2 infection (aHR 0.60 [0.43-0.84]). Sensitivity of self-reported COVID-19 vaccination status was 82% (80-85%) overall, and 98% (97-99%) among those referencing vaccination records; specificity was 87% (86-89%).

**Interpretation:** Participants’ stated willingness to receive COVID-19 vaccination was an imperfect predictor of real-world vaccine receipt. Improving messaging about the importance of COVID-19 vaccination, regardless of previous SARS-CoV-2 infection status, may improve vaccine uptake among populations who express hesitancy to initiate vaccination.

**RESEARCH IN CONTEXT:** *Evidence before this study:* We searched PubMed and medRχiv for variations and combinations of the terms “vaccine hesitancy”, “vaccine confidence”, “vaccine uptake”, “COVID-19”, and “SARS-CoV-2” to identify original research articles published by March 8, 2022. The majority of screened articles were cross-sectional surveys conducted prior to or after implementation of COVID-19 vaccines to assess trends or predictors of participant-reported COVID-19 vaccine hesitancy. While some studies included random population-based samples, many were conducted within subgroups like health care professionals, parents of school aged children, or college students. Evidence about the association between COVID-19 vaccine intentions and subsequent vaccine uptake remains scarce. Three observational studies quantified associations between willingness to receive COVID-19 vaccination and subsequent initiation of vaccination; however, in these studies, follow-up time was limited to the period prior to widespread availability of COVID-19 vaccination or initiation of vaccine mandates in workplaces, schools, and other public places. Therefore, it was unclear whether remaining unvaccinated at follow-up in these studies was a choice or a consequence of the lack of universal access to COVID-19 vaccines. Additionally, most efforts to identify subsequent vaccine uptake relied on self-reported vaccination status, which may be subject to reporting or interviewer bias. We also searched PubMed and medRχiv with variations and combinations of the terms “self-reported”, “vaccination”, “accuracy”, and “COVID-19” and did not discover any articles validating self-reported COVID-19 vaccination status against immunization registry data; whereas, such studies were available for other vaccine-preventable pathogens including influenza, *Streptococcus pneumoniae*, and human papillomavirus.

*Added value of this study:* We linked data collected through an ongoing case-control study and a comprehensive state-wide immunization registry to evaluate the association between COVID-19 vaccination intention and subsequent uptake. We also assessed the reliability of self-reported COVID-19 vaccination status by linking participant records with a state-wide immunization registry. We are not aware of another published study assessing predictors of COVID-19 vaccine uptake spanning over 7 months of age-eligible follow-up time and adjudicating the use of self-reported COVID-19 vaccination status. We found that expressing hesitancy to receive COVID-19 vaccination was associated with lower adjusted hazards of subsequent vaccine uptake as compared with expressing willingness to receive vaccination (aHR: 0.49; 95% CI: 0.32-0.76), although uptake was also suboptimal among individuals who expressed willingness (45%). Participants from lower income households or who had recently tested positive for SARS-CoV-2 were slower to initiate vaccination than from higher income households or who had recently tested negative. People who were pregnant and initially deferred vaccination were faster to receive vaccination than participants who did not cite pregnancy as a reason for refusal. Upon assessing the accuracy of self-reported vaccination status, we found referencing a vaccination card or another calendar reference source improved sensitivity of self-reported vaccination status.

*Implications of all available evidence:* We provide an evaluation of predictors of COVID-19 vaccine uptake and assess the validity of self-reported COVID-19 vaccination status in comparison with a state-wide immunization registry. We identified that self-reported vaccination intent was a strong but imperfect predictor of subsequent vaccine initiation. However, no single reason for participants to express vaccine hesitancy predicted their likelihood of eventual vaccine receipt. As such, public health campaigns addressing multiple factors underlying vaccine hesitancy including those correcting sources of misinformation, and allaying concerns about short- or long-term side effects and vaccine safety remain important tools to improve acceptance in hesitant populations. Future studies reliant on the use of self-reported COVID-19 vaccination status should strive to utilize additional reference sources like COVID-19 vaccination cards or vaccination registries to reduce misclassification of vaccination status.

## Background

Suboptimal uptake of COVID-19 vaccination among eligible individuals in the United States (US) and other settings has contributed to preventable COVID-19 cases, hospitalizations, and deaths [1]. Addressing barriers to timely vaccination against COVID-19 is thus a priority to mitigate disease burden. While surveys have provided an important tool for assessing vaccine hesitancy and acceptance across differing communities, alignment between participants’ self-reported vaccine intentions and real-world receipt of vaccination is not well understood [2,3]. Understanding barriers and facilitators of COVID-19 vaccine receipt among individuals who express hesitancy, or that preclude vaccine access among individuals who express willingness, could support efforts to maximize vaccine uptake.

The State of California first made COVID-19 vaccines available to health care workers in November 2020; by April 19, 2021, eligibility for COVID-19 vaccination expanded to all California residents aged 16 years and older [4,5]. Healthcare providers administering COVID-19 vaccines in California are required to report all doses administered to local or state-level public health authorities, enabling comprehensive tracking of vaccine uptake within the state’s population via the state-wide immunization registry As of December 5, 2021, 28.5 million (73%) of California’s 39.2 million residents were recorded as having received ≥1 doses of any COVID-19 vaccine within the state [6].

As part of efforts to inform vaccine rollout in California, the California Department of Public Health (CDPH) collected data on willingness to receive COVID-19 vaccines on a continuous basis among participants in a test-negative design case-control study through the period of COVID-19 vaccine rollout [4,7,8]. To understand the relationship between participants’ self-reported vaccine intentions and real-world vaccine uptake, we cross-referenced data from study participants and the state-wide immunization registry to compare COVID-19 vaccine receipt among individuals who expressed hesitancy or willingness to be vaccinated. To further inform uses of self-reported vaccination in research studies, we assessed the accuracy of participants’ self-reported COVID-19 vaccination status in comparison with registry-based documentation of COVID-19 vaccination.

## Methods

### Study Population

This analysis used data from participants enrolled in the California COVID-19 Case-Control study, which was undertaken to evaluate risk factors for SARS-CoV-2 infection within the state. Survey methodology has been described elsewhere [7,8]. In brief, enrolled participants were individuals who received a molecular SARS-CoV-2 test in California and had no known history of a previous SARS-CoV-2 infection (molecular, antigen, serological). Throughout the study period, all SARS-CoV-2 molecular tests occurring in California were required to be reported to the state health department. Potential case (SARS-CoV-2 test positive) and control (SARS-CoV-2 test negative) participants were individually matched on age, sex, region of residence in California, and date of SARS-CoV-2 test result report. Each day throughout the study period, trained interviewers administered a telephone-based structured questionnaire in English and Spanish to randomly selected California residents from among all those with a confirmatory SARS-CoV-2 test result posted to the California Reportable Disease Registry in the preceding 48 hours. Interviewers documented sociodemographic characteristics and self-reported COVID-19 vaccination status; among participants who reported receiving one or more doses of a COVID-19 vaccine product, interviewers recorded the self-reported date(s) of receipt and manufacturer of each dose. Participants were encouraged to refer to their COVID-19 vaccination card or another recall aid (e.g., e-mail, text message, calendar reminder, and/or diary entry) when providing vaccine history. Participants included in this analysis were ≥5 years of age and enrolled from 19 April 2021 (when all Californians aged ≥16 years became eligible to receive COVID-19 vaccines) to 5 December 2021, when the database was locked for linkage to the state-wide immunization registry.

### State-wide Immunization Registry

The state of California tracks receipt of COVID-19 vaccination to monitor trends in vaccination (e.g., at a geographic level or by antigen), identify potential gaps in vaccination coverage, and inform public health efforts to improve immunization. Healthcare providers in 49 of 58 California counties (collectively accounting for 87% of California’s population) submit data on vaccine administration directly to the state-wide immunization registry on all COVID-19 vaccine doses administered. In the remaining nine counties, data are linked to the state-wide immunization registry from local-level registries. The San Diego Immunization Registry (SDIR) collects data from providers in San Diego County, while the Healthy Futures (HF) Immunization Registry collects data from providers in the remaining eight counties (Alpine, Amador, Calaveras, Mariposa, Merced, San Joaquin, Stanislaus, and Tuolumne); the state-wide immunization registry receives data from these regional IISs rather than by direct notification from healthcare providers. Reports on >90% of doses administered within the state of California are received by the state-wide immunization registry within one day of the vaccine administration date. However, some vaccination dose submissions may be less timely, such as those from mass vaccination clinics or those that are manually entered into the state-wide immunization registry. The state-wide immunization registry intends to capture all COVID-19 vaccinations occurring within the state of California.

### Inclusion and exclusion criteria

The time-to-vaccination analysis was restricted to participants enrolled from 19 April 2021 onward. Eligibility to receive vaccination was defined by age, per date of the United States Health and Human Services recommendation for a particular age group to receive COVID-19 vaccination. Participants aged 5-64 years who indicated they had not yet received any doses of a COVID-19 vaccine at the time of enrollment were eligible for inclusion in this sub-study, including primary analysis of factors predicting vaccine receipt (**Figure 1**). Participants aged 0-4 years were excluded from this analysis due to their ineligibility for COVID-19 vaccination throughout the study period; individuals aged ≥65 years were excluded because differing dates of vaccine eligibility for residents of long-term care facilities or other adults aged ≥65 years precluded reliable measurement of the time from when individuals became eligible for COVID-19 vaccines to when they received an initial dose. We further excluded participants who self-reported medical contraindications for receiving COVID-19 vaccines.

**Figure 1.**
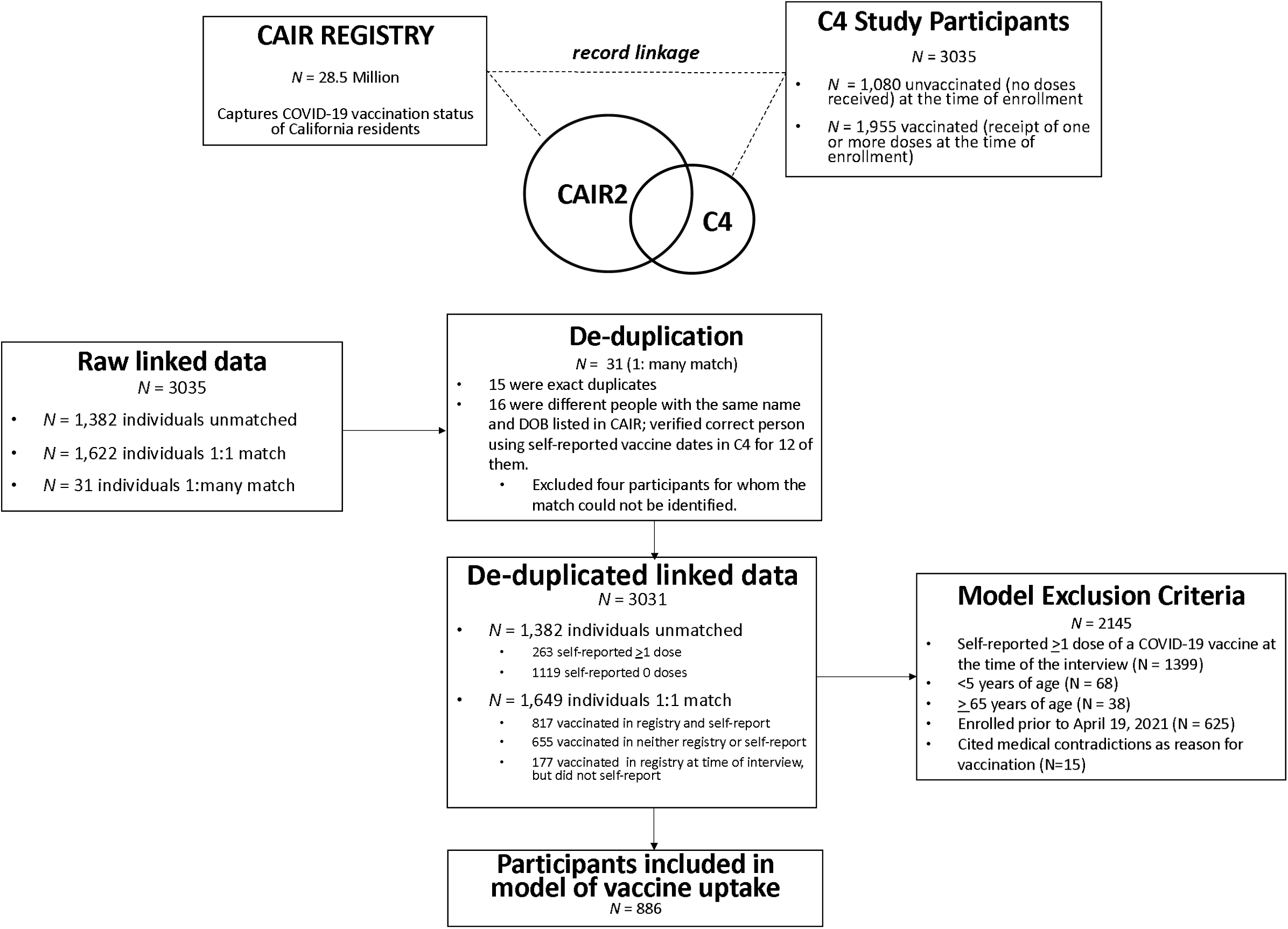
Flow chart of participants included in CAIR, C4 data, and ultimately the analytic data set.

### Record Linkage

We linked participant records across the study and immunization registry using a previously-described probabilistic framework [9]. We first identified records of vaccine doses administered among all study participants by searching for exact or deterministic matches on zip code of residence and date of birth, and fuzzy matches on first and last name (standardizing text fields by removing uppercase letters, spaces, and special characters). We undertook manual review of records if one participant was matched to multiple vaccine records, and for all participants with match assignment probabilities valued between 0.5 and 0.9525. Participants in the study were considered to have no documented receipt of COVID-19 vaccine doses if this manual record review identified no prospective matches with probabilities <0.9525 explainable by data entry errors.

### Statistical Analysis

Our primary outcome of interest was the time from age-eligibility of COVID-19 vaccination to COVID-19 vaccine initiation as recorded in the immunization registry or censoring, if no COVID-19 vaccine doses were received. Participants aged ≥16 years, ≥11 years, or ≥5 years were considered age-eligible for COVID-19 vaccination on April 19, May 10, and October 29, 2021, respectively [5]. Participants who were vaccinated prior to the date of eligibility (*n*=11) were assigned an observation time of 1 day.

We used Cox proportional hazards models to estimate adjusted hazard ratios (aHR) of vaccine uptake throughout the study period. The primary exposure of interest was self-reported intention to receive COVID-19 vaccination. Models adjusted for age, race/ethnicity, sex, annual household income, region of residence within California (**Table S1**), SARS-CoV-2 test result status at the time of enrollment in the study, self-reported comorbid conditions, and self-reported uptake of/adherence to public health mitigation measures including use of face masks and physical distancing. To account for differences in the study population who remained unvaccinated throughout the study period, participants were compared within regression strata defined by the calendar month of participation in the study. We verified the proportional hazards assumption by testing for slopes in Schoenfeld residuals [10].We then repeated these analyses in subgroups of participants according to their stated intentions of receiving COVID-19 vaccination. As secondary analyses, we assessed differences in time to initiate COVID-19 vaccination according to participants’ stated reasons for vaccine refusal or hesitancy using Cox proportional hazards models, restricted to participants who stated they were unwilling or hesitant to receive vaccination.

We also sought to validate participants’ self-reported vaccination status using the immunization registry. Participants were each categorized into four mutually exclusive categories according to alignment of their self-reported vaccination status and linked data from the immunization registry: self-reported vaccinated with match in immunization registry (A), self-reported vaccinated without match in immunization registry (B), self-reported unvaccinated and match in immunization registry (C), or self-reported unvaccinated and without immunization registry match (D). Vaccination status in the immunization registry was recoded to match the vaccination status of a participant at the time of their telephone interview. Sensitivity, specificity, positive predictive value (PPV), and negative predictive value (NPV) of self-reported vaccination status as compared with registry-documented vaccination status (treated as the “gold standard”) were calculated with accompanying 95% confidence intervals via bootstrap resampling. We additionally stratified these calculations by SARS-CoV-2 test result, enrollment period in the study, use of a recall aid at the time of study participation, age, and region. As linkage analyses did not entail measurement of time from vaccine eligibility to vaccine receipt, participants of all ages were included in these analyses. As a sensitivity analysis, we conducted a quantitative bias analysis to assess the extent to which vaccine effectiveness estimates derived from self-reported vaccination status in epidemiologic data sets may be biased due to differential sensitivity and specificity between cases and controls.

All analyses were performed using R (version 3.6.1; R Foundation for Statistical Computing). The probabilistic match approach was completed using the *RecordLinkage* package [11]. We used the *survival* package for time-to-event analyses [12]. We used the *episensr* package for quantitative bias analysis.

### Ethics

All participants provided oral informed consent. For minors (<18 years of age), assent from participant and informed consent from a parent/guardian were both required. The study protocol was approved by the State of California, Health and Human Services Agency, Committee for the Protection of Human Subjects (Project Number: 2021-034).

## Findings

Among 3,035 individuals enrolled between 24 February 2021 – 5 December 2021 and who self-reported their COVID-19 vaccination status, 54% (1622/3035) matched with a single record in the immunization registry, 45.5% (1382/3035) did not have a record of COVID-19 vaccination, and 1% (31/3035) individuals matched with multiple records **(Figure 1**). Upon de-duplication of the 31 records with a one-to-many match in the state-wide immunization registry, four records were excluded due to the inability to identify a correct match. Ultimately, 3031 participants were included in the assessment of accuracy of self-reported COVID-19 vaccination status, among whom 35.6% (1080) self-reported having already received ≥1 dose of COVID-19 vaccine at time of study enrollment. The majority of participants were adults, aged over 18 years (86.9%; 2635/3031), and participants were enrolled equally across urban and rural regions in California (**Table S2**).

For the primary analysis we included a total of 886 individuals who met the inclusion criteria outlined above (**Figure 1**). We found 423 (47.7%) were willing to, 185 (20.8%) were unsure, and 278 (31.3%) were unwilling to receive COVID-19 vaccination (**Table 1; Table S3; Figure 2**). Among the 278 individuals who were unwilling to receive COVID-19 vaccination, 13% (37/278) matched with a vaccine record by 5 December 2021. Similarly, among the 185 individuals who were unsure about receiving COVID-19 vaccination, 22% (41/185) matched with a vaccine record. Of participants who were willing to receive COVID-19 vaccination, 46% (194/423) matched with a vaccine record as of 5 December 2021.

**Table 1.**
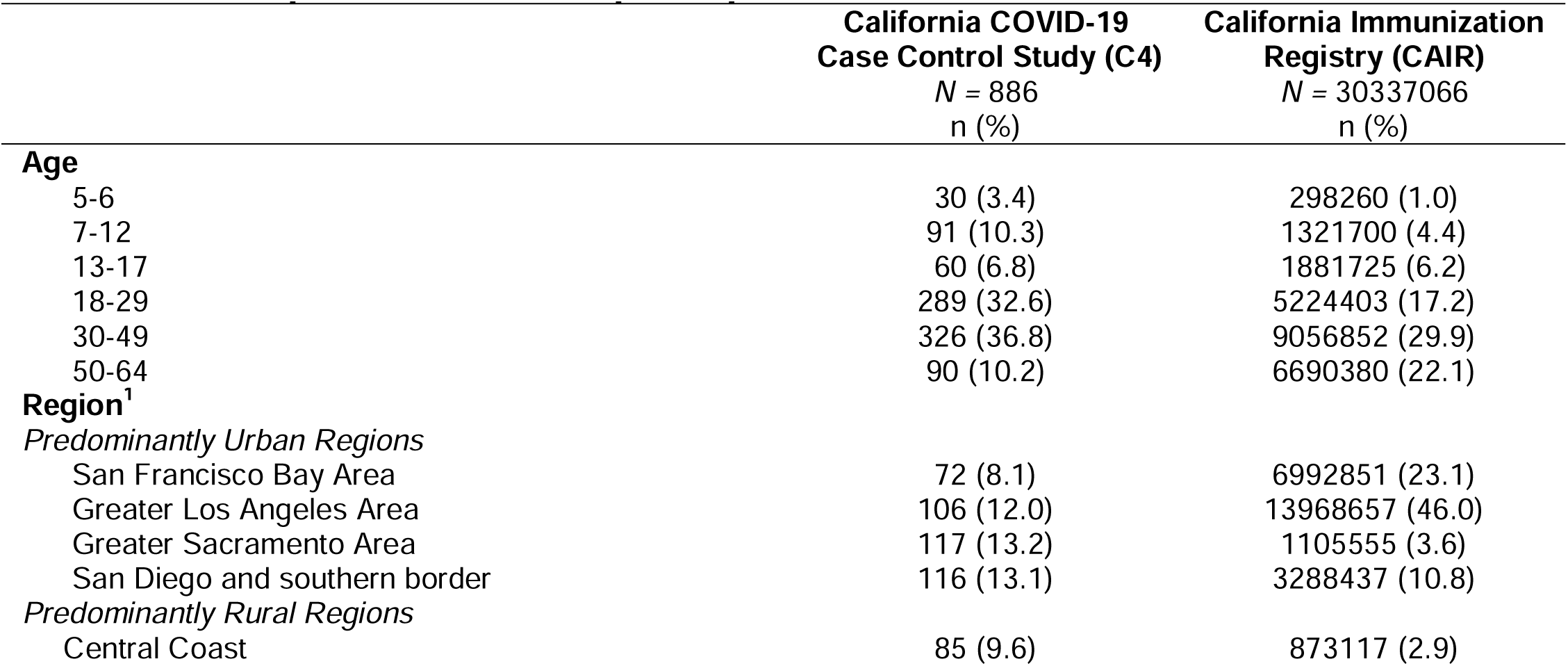
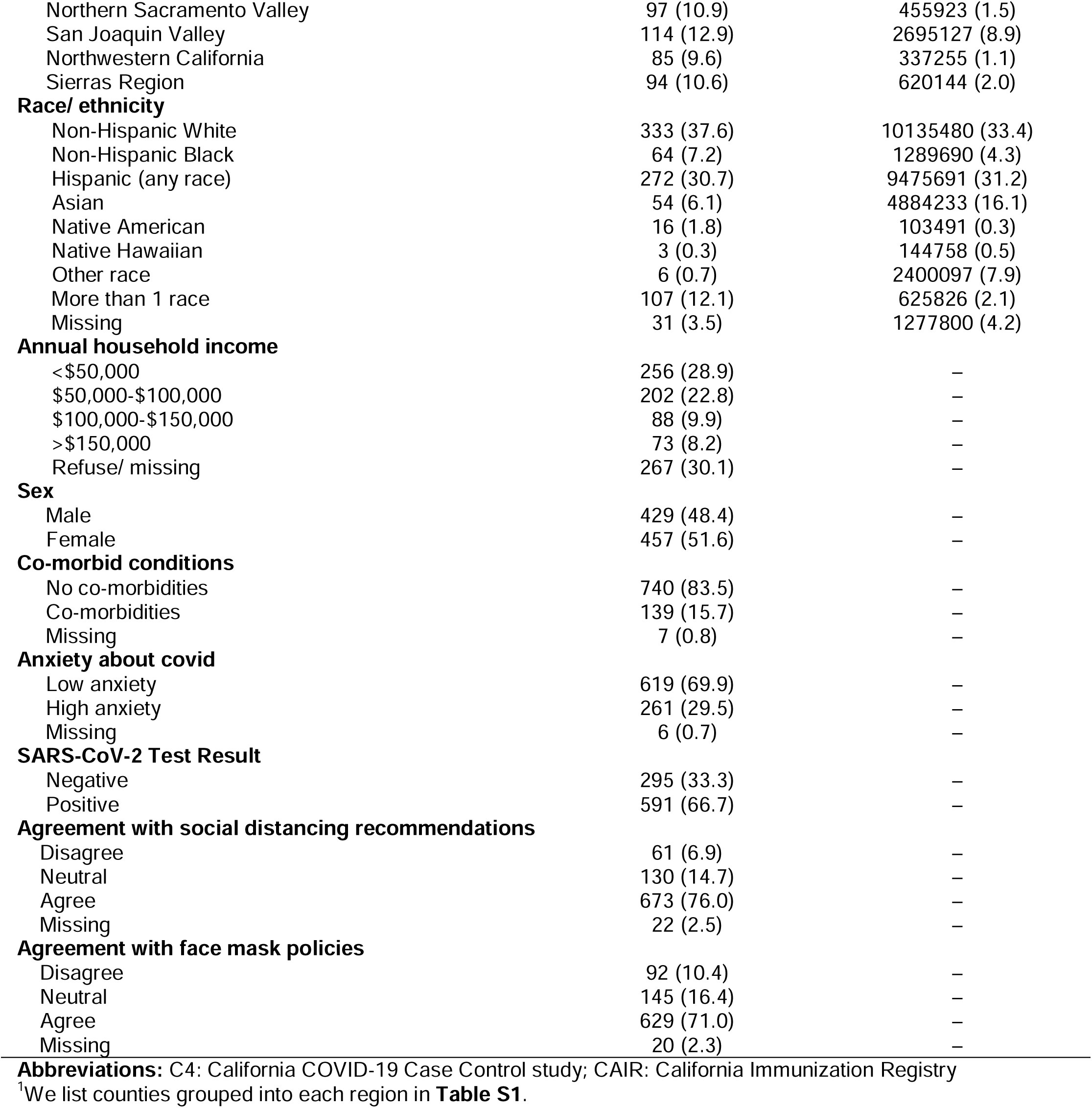
Descriptive attributes of participants included in CAIR and C4 data.

**Figure 2.**
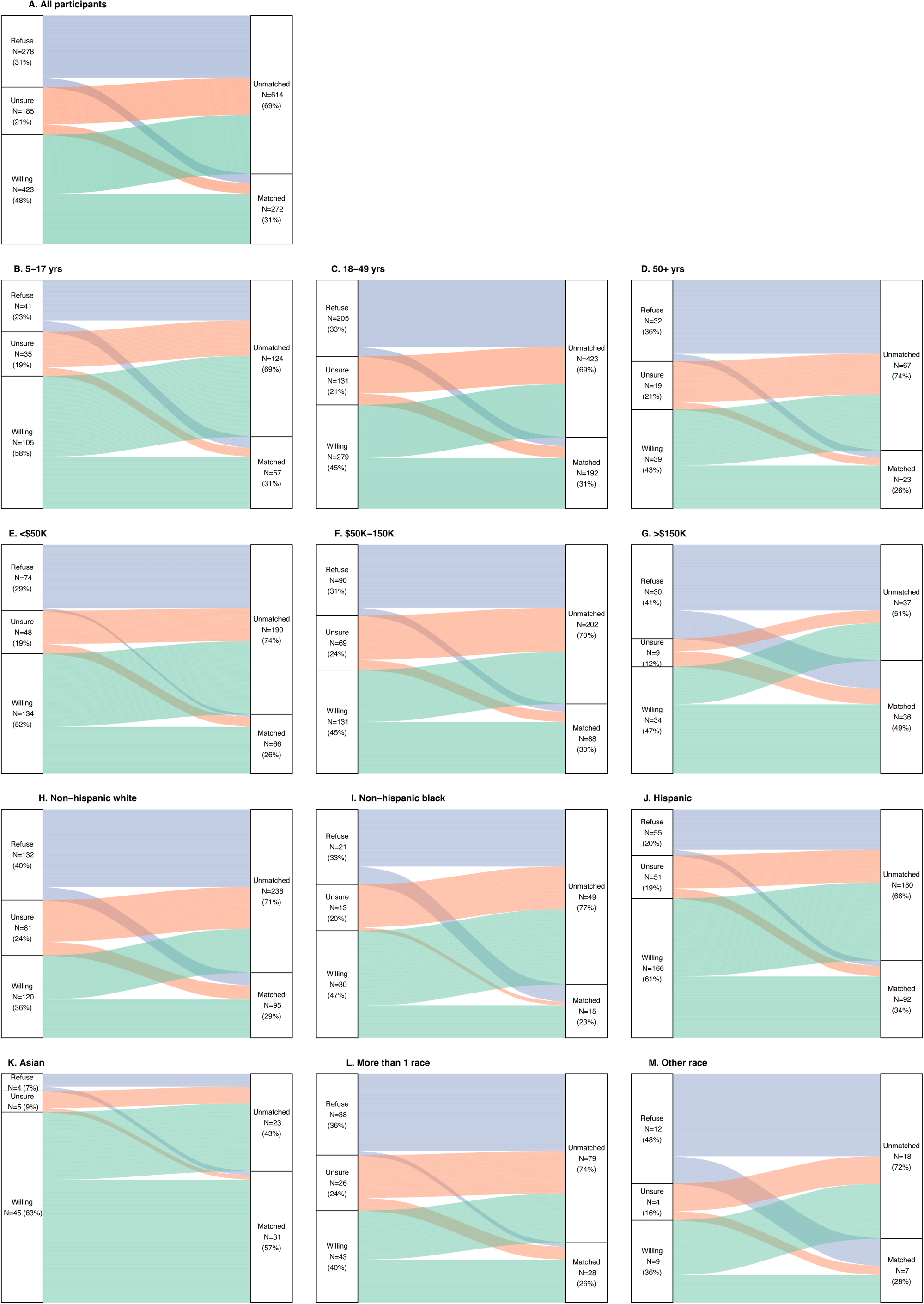
Stated vaccine acceptance and subsequent vaccine uptake among study participants. (attached)

Adjusted hazard ratio estimates indicated longer time to COVID-19 vaccine uptake among participants who stated they were unsure (aHR: 0.49 [95% CI: 0.32-0.76]) or unwilling (aHR: 0.21 [0.12-0.36]) to initiate COVID-19 vaccination, as compared with participants expressing willingness to be vaccinated (**Table 2; Figure S1**). The adjusted hazard ratio of vaccine uptake comparing female with male participants was 1.56 (95% CI: 1.12-2.17). Children aged 5-12 (aHR: 2.37 [1.30-4.33]) and teenagers aged 13-17 (aHR: 2.09 [1.13-3.88]) experienced shorter time to receive vaccination than young adults aged 18-29. Participants from households with an annual income greater than $150,000 had 3.30 (95% CI: 2.02-5.39) times higher adjusted hazards of receiving a COVID-19 vaccine than participants from households with annual income under $50,000. Case participants (SARS-CoV-2 test positive) experienced longer time to vaccinate (aHR: 0.60 [0.43-0.84]) than control participants (SARS-CoV-2 test negative). Time to vaccination was shorter among participants reporting co-morbidities or immunocompromising conditions as compared to (aHR: 1.54 [1.01-2.36]) those without health conditions. We did not observe statistically significant differences in the time to uptake of COVID-19 vaccination by race/ethnicity, region of residence, or self-reported anxiety about the pandemic or adherence to COVID-19 mitigation measures including use of face masks or physical distancing.

**Table 2.** Predictors of time to vaccine uptake among participants (*N* = 886) who self-reported that they had not yet received any doses of a SARS-CoV-2 vaccine at the time of the C4 interview.

| Participant characteristic | All participants |  |  |
| --- | --- | --- | --- |
|  | Proportion vaccinated<br>Number vaccinated/Total (%) | HR (95% CI) |  |
|  |  | Unadjusted | Adjusted |
| Vaccine willingness during C4 interview |  |  |  |
| Willing to get the COVID-19 vaccine | 194/423 (45.9) | ref. | ref. |
| Unsure about the COVID-19 vaccine | 41/185 (22.2) | 0.40 (0.29, 0.56) | 0.49 (0.32,0.76) |
| Unwilling to get the COVID-19 vaccine | 37/278 (13.3) | 0.23 (0.16,0.33) | 0.21 (0.12,0.36) |
| Age of participant |  |  |  |
| 5-12 | 38/121 (31.4) | 5.24 (3.40, 8.09) | 2.37 (1.30,4.33) |
| 13-17 | 19/60 (31.7) | 1.30 (0.79, 2.14) | 2.09 (1.13,3.88) |
| 18-29 | 97/289 (33.6) | ref. | ref. |
| 30-49 | 95/326 (29.1) | 0.87 (0.55, 1.16) | 0.96 (0.65,1.41) |
| 50-64 | 23/90 (25.6) | 0.74 (0.47, 1.18) | 0.71 (0.35,1.44) |
| Region<sup>1</sup> |  |  |  |
| San Francisco Bay Area | 36/72 (50.0) | ref. | ref. |
| Greater Los Angeles Area | 34/106 (32.1) | 0.49 (0.31, 0.79) | 0.71 (0.38,1.34) |
| Greater Sacramento Area | 36/117 (30.8) | 0.52 (0.33, 0.83) | 0.68 (0.37,1.26) |
| San Diego and southern border | 39/116 (33.6) | 0.66 (0.42, 1.05) | 1.09 (0.59,2.02) |
| Central Coast | 24/85 (28.2) | 0.38 (0.22, 0.65) | 0.51 (0.26,1.04) |
| Northern Sacramento Valley | 26/97 (26.8) | 0.40 (0.24, 0.67) | 0.81 (0.38,1.71) |
| San Joaquin Valley | 32/114 (28.1) | 0.51 (0.32, 0.82) | 0.62 (0.32,1.17) |
| Northwestern California | 19/85 (22.4) | 0.37 (0.21, 0.64) | 0.70 (0.34,1.45) |
| Sierras Region | 26/94 (27.7) | 0.43 (0.26, 0.72) | 0.85 (0.40,1.80) |
| Annual household income |  |  |  |
| <\$50,000 | 66/256 (25.8) | ref. | ref. |
| \$50,000-\$100,000 | 57/202 (28.2) | 1.07 (0.75, 1.54) | 1.71 (1.14,2.55) |
| \$100,000-\$150,000 | 31/88 (35.2) | 1.52 (0.99, 2.33) | 1.76 (1.06,2.91) |
| >\$150,000 | 36/73 (49.3) | 2.22 (1.46, 3.37) | 3.30 (2.02,5.39) |
| Race/ethnicity |  |  |  |
| Non-Hispanic white | 97/339 (23.4) | ref. | ref. |
| Asian | 31/54 (57.4) | 2.79 (1.86, 4.20) | 1.67 (0.91,3.08) |
| Black | 15/64 (23.4) | 0.73 (0.42, 1.25) | 1.24 (0.53,2.88) |
| Hispanic | 92/272 (33.8) | 1.19 (0.90, 1.59) | 1.50 (0.98,2.28) |
| More than 1 race | 28/107 (26.2) | 0.96 (0.63, 1.46) | 0.96 (0.54,1.71) |
| Other | 5/19 (26.3) | 0.91 (0.33, 2.49) | 0.76 (0.21,2.67) |
| Sex |  |  |  |
| Male | 116/429 (27.0) | ref. | ref. |
| Female | 156/457 (34.1) | 1.28 (1.01, 1.63) | 1.56 (1.12,2.17) |
| Co-morbid conditions |  |  |  |
| No co-morbidities | 227/740 (30.7) | ref. | ref. |
| Co-morbidities | 43/139 (30.9) | 0.93 (0.67, 1.28) | 1.54 (1.01,2.36) |
| Anxiety about covid |  |  |  |
| Low anxiety | 176/ 619 (28.4) | ref. | ref. |
| High anxiety | 93/261 (35.6) | 1.37 (1.07, 1.76) | 1.05 (0.75,1.49) |
| SARS-CoV-2 Test Result |  |  |  |
| Negative | 103/295 (34.9) | ref. | ref. |
| Positive | 169/591 (28.6) | 0.68 (0.53, 0.87) | 0.60 (0.43,0.84) |
| Agreement with social distancing recommendations |  |  |  |
| Disagree | 9/61 (14.8) | ref. | ref. |
| Neutral | 22/130 (16.9) | 1.33 (0.63, 2.80) | 1.50 (0.40,5.69) |
| Agree | 232/673 (34.5) | 2.52 (1.34, 4.76) | 2.31 (0.66,8.15) |
| Agreement with face mask policies |  |  |  |
| Disagree | 12/92 (13.0) | ref. | ref. |
| Neutral | 33/145 (22.8) | 2.10 (1.10, 3.98) | 1.16 (0.43,3.12) |
| Agree | 219/629 (34.8) | 3.06 (1.75, 5.37) | 1.04 (0.41,2.65) |

**Abbreviations:** C4: California COVID-19 Case Control study; ref: reference category; HR: Hazard Ratio
<sup>1</sup>We list counties grouped into each region in **Table S1**.

Among the subgroup of participants who stated they were unwilling or unsure about receiving COVID-19 vaccination, younger participants aged 5-12 years (aHR: 14.19 [2.15-93.33]) and 13-17 years (aHR: 3.98 [1.22-12.94]) experienced shorter time to vaccinate compared with participants aged 18-29 years (**Table 3**). Within the subgroup of participants who indicated they were willing to receive COVID-19 vaccination, there were not significant differences in the time to vaccine uptake by age, although point estimates suggested higher uptake among children aged under 18 years (aHR: 1.74 and 1.70 for those aged 5-12 years and 13-17 years, respectively, as compared with 18-29 years). The adjusted hazard ratio of subsequent vaccination among higher income (>$150,000) households as compared with lower income (<$50,000) households among the unwilling or unsure and willing were 5.53 (1.85-16.57) and 2.69 (1.47-4.93), respectively. Time to vaccine uptake was longer among participants who had recently tested positive for SARS-CoV-2 (aHR: 0.23 [0.11-0.47]) if they indicated they were unwilling or hesitant to initiate COVID-19 vaccination; however, this effect was not apparent among the SARS-CoV-2 positive participants who indicated willingness to initiate vaccination (aHR: 0.92 [0.60-1.40]). Differences in the time to vaccine uptake were not apparent within subgroups of participants who were unwilling or hesitant to receive vaccination according to region, race/ethnicity, sex, presence of co-morbidities, self-reported anxiety about the pandemic or adherence to COVID-19 mitigation measures including use of face masks or physical distancing. Leading reasons for reporting as unsure or unwilling to receive vaccine were concerns about COVID-19 vaccine safety and/or side effects (43%; 199/653), wanted to wait for more research or learn more about COVID-19 vaccines (36%; 165/463), and/or had ideological reasons (21%; 98/463) associated with adjusted hazard ratio estimates of subsequent vaccination 0.91 (0.48, 1.70), 1.10 (0.58, 2.11), and 0.69 (0.24-2.01), respectively. Time to vaccine uptake was longer among participants who indicated that COVID-19 vaccination should be their personal choice (16%, 76/463) as compared to participants who did not cite this reason (aHR: 0.62 [0.18-2.07]), although this association was not statistically significant. None of the eight participants who cited religious objections as a reason to be unsure or unwilling to receive vaccine subsequently received vaccination as of 5 December 2021. Participants who were pregnant at the time of the telephone interview experienced shorter time to vaccine uptake (aHR: 4.19 [1.41-12.41]) than those who did not cite pregnancy as a reason for not receiving vaccination.

**Table 3.**
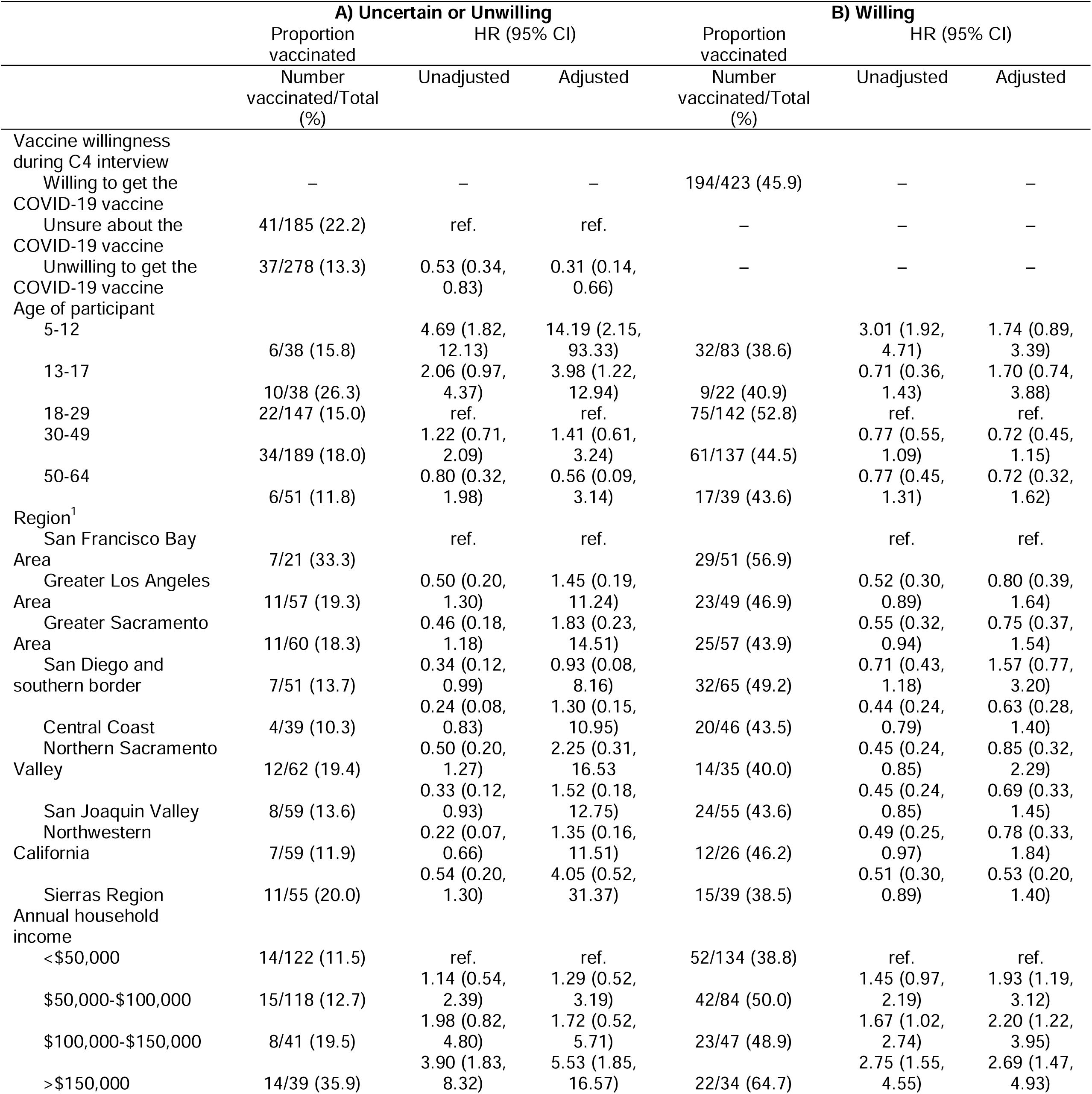
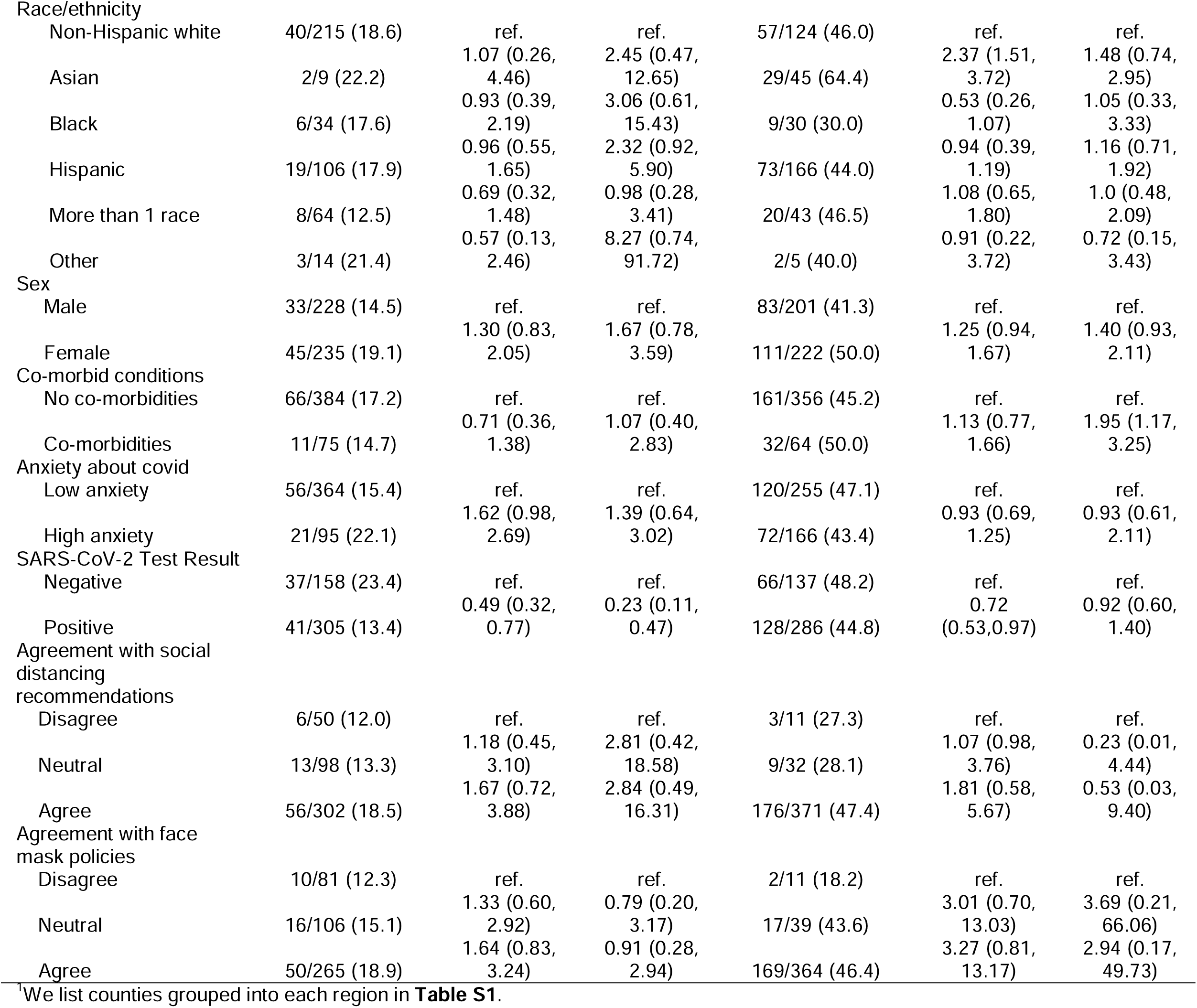
Predictors of time to vaccine uptake restricted to participants who were A) unwilling or hesitant participants or B) willing to receive SARS-CoV-2 vaccination during the C4 telephone interview.

The sensitivity and specificity of self-reporting receipt of one or more doses of COVID-19 vaccine was 82% (95% CI: 80–85%) and 87% (86–89%), respectively, in comparison to vaccine doses recorded in the immunization registry at the time of the telephone interview **(Table 5)**. The positive predictive value (PPV) and negative predictive value (NPV) of participant-reported vaccination status were 76% (73–78%) and 91% (90–92%), respectively. Sensitivity of self-reported vaccination status was significantly higher among participants who referenced a recall aid; sensitivity was 98% (97-99%) among participants who referenced their vaccination card, 92% (86-96%) among participants who referenced another recall aid (e.g., e-mail, text message, calendar reminder, and/or diary entry for their vaccine appointment), and 45% (38-52%) among participants who did not reference a recall aid. Sensitivity (86% [82-90%] vs. 80% [77-83%]) and specificity (94% [92-95%] vs. 78% [75-81%]) of self-reported vaccination status was significantly higher among cases compared with controls. In a quantitative bias analysis, differential misclassification of self-reported vaccination status by SARS-CoV-2 test result resulted in non-significant overestimates of COVID-19 vaccine effectiveness (**Table S4**). No differences in sensitivity and specificity were apparent within strata of SARS-CoV-2 test result and use of a recall aid; thus, accounting for use of a recall aid results in non-differential misclassification of vaccination status, resulting in underestimates of vaccine effectiveness.

**Table 5.** Comparison of vaccination status defined by the California Immunization Registry (CAIR) and vaccination self-report using data from the California COVID-19 case-control study. Characteristics of participants included in this analysis are listed in **Table S2**.

|  | CAIR-Vaccinated<br>N | CAIR-Unvaccinated<br>N | Sensitivity<br>(95% CI) | Specificity<br>(95% CI) | PPV<br>(95% CI) | NPV<br>(95% CI) |
| --- | --- | --- | --- | --- | --- | --- |
| **A. All participants** |  |  |  |  |  |  |
| Self-report Vaccinated | 817 | 263 | 0.82 (0.80, 0.85) | – | 0.76 (0.73, 0.78) | – |
| Self-report Unvaccinated | 177 | 1,774 | – | 0.87 (0.86, 0.89) | – | 0.91 (0.90, 0.92) |
| **B. Use of Recall Aid** |  |  |  |  |  |  |
| Self-report vaccinated<br>referenced vaccine card | 577 | 180 | 0.98 (0.97, 0.99) | – | 0.76 (0.73, 0.78) | – |
| Self-report vaccinated with<br>another recall aid (ex. e-mail<br>or calendar) | 137 | 51 | 0.92 (0.86, 0.96) | – | 0.73 (0.66, 0.79) | – |
| Self-report vaccinated<br>without recall aid | 98 | 32 | 0.45 (0.38, 0.52) | – | 0.75 (0.67, 0.83) | – |
| **C. SARS-CoV-2 Positive** |  |  |  |  |  |  |
| Self-report Vaccinated | 299 | 74 | 0.86 (0.82, 0.90) | – | 0.80 (0.76, 0.85) | – |
| Self-report Unvaccinated | 48 | 1098 | – | 0.94 (0.92, 0.95) | – | 0.96 (0.94, 0.97) |
| **D. SARS-CoV-2 Negative** |  |  |  |  |  |  |
| Self-report Vaccinated | 518 | 189 | 0.80 (0.77, 0.83) | – | 0.73 (0.70, 0.76) | – |
| Self-report Unvaccinated | 129 | 676 | – | 0.78 (0.75, 0.81) | – | 0.84 (0.81, 0.86) |
| **E. Test Status & Recall Aid** |  |  |  |  |  |  |
| *SARS-CoV-2 Positive* |  |  |  |  |  |  |
| Self-report vaccinated<br>referenced vaccine card | 270 | 65 | 0.97 (0.95, 0.99) | – | 0.81 (0.76, 0.85) | – |
| Self-report vaccinated<br>without recall aid | 27 | 9 | 0.48 (0.35, 0.62) | – | 0.75 (0.58, 0.88) | – |
| *SARS-CoV-2 Negative* |  |  |  |  |  |  |
| Self-report vaccinated<br>referenced vaccine card | 444 | 166 | 0.97 (0.95, 0.98) | – | 0.73 (0.69, 0.76) | – |
| Self-report vaccinated<br>without recall aid | 71 | 23 | 0.44 (0.36, 0.51) | – | 0.76 (0.66, 0.84) | – |

**Abbreviations:** CAIR: California Immunization Registry; PPV: positive predictive value; NPV: negative predictive value

Sensitivity and specificity of self-reported vaccination status were highest among individuals vaccinated during the early stages (February – April 2021) of vaccine roll-out in California (93% [87–96%] and 94% [92–96%], respectively); among those vaccinated during August – December 2021, sensitivity and specificity of self-reported vaccination status were 78% (73-–82%) and 81% (77–85%), respectively (**Table 6**). While sensitivity of self-reported vaccination status was significantly lower among children (<18 years of age) than adults (≥18 years of age), specificity was higher amongst children than adults. Differences in the accuracy of self-reported vaccination status were not apparent in analyses stratified by urban and rural regions of the state. Sensitivity analyses estimated agreement between self-reported COVID-19 vaccine manufacturer and dates of initiating COVID-19 vaccination upon comparison of self-report and registry-based documentation (**Table S5; Table S6**).

**Table 6.** Comparison of vaccination status defined by the California Immunization Registry (CAIR) and vaccination self-report, stratified by A) time period of self-report in the C4 study, B) age of participant, C) California region. Characteristics of participants included in this analysis are listed in **Table S2**.

|  | CAIR-Vaccinated<br>N | CAIR -<br>Unvaccinated<br>N | Sensitivity<br>(95% CI) | Specificity<br>(95% CI) | PPV<br>(95% CI) | NPV<br>(95% CI) |
| --- | --- | --- | --- | --- | --- | --- |
| **A. Time period of enrollment** |  |  |  |  |  |  |
| **February 21 – April 8, 2021** |  |  |  |  |  |  |
| Self-report Vaccinated | 128 | 38 | 0.93 (0.87, 0.96) | – | 0.77 (0.70, 0.83) | – |
| Self-report Unvaccinated | 10 | 608 | – | 0.94 (0.92, 0.96) | – | 0.98 (0.97, 0.99) |
| **April 9 – May 23, 2021** |  |  |  |  |  |  |
| Self-report Vaccinated | 184 | 82 | 0.88 (0.82, 0.92) | – | 0.69 (0.63, 0.75) | – |
| Self-report Unvaccinated | 26 | 447 | – | 0.84 (0.81, 0.87) | – | 0.95 (0.92, 0.96) |
| **May 24 – August 12, 2021** |  |  |  |  |  |  |
| Self-report Vaccinated | 215 | 72 | 0.78 (0.73, 0.83) | – | 0.75 (0.69, 0.80) | – |
| Self-report Unvaccinated | 59 | 412 | – | 0.85 (0.82, 0.88) | – | 0.87 (0.84, 0.90) |
| **August 13 – December 5, 2021** |  |  |  |  |  |  |
| Self-report Vaccinated | 290 | 71 | 0.78 (0.73, 0.82) | – | 0.80 (0.76, 0.84) | – |
| Self-report Unvaccinated | 82 | 307 | – | 0.81 (0.77, 0.85) | – | 0.79 (0.75, 0.83) |
| **B. Age of Study Participant** |  |  |  |  |  |  |
| **≤ 12 years old** |  |  |  |  |  |  |
| Self-report Vaccinated | 5 | 1 | 0.62 (0.24, 0.91) | – | 0.83 (0.36, 1.00) | – |
| Self-report Unvaccinated | 3 | 231 | – | 1.00 (0.98, 1.00) | – | 0.99 (0.96, 1.00) |
| **13-17 years old** |  |  |  |  |  |  |
| Self-report Vaccinated | 19 | 1 | 0.61 (0.42, 0.78) | – | 0.95 (0.75, 1.00) | – |
| Self-report Unvaccinated | 12 | 125 | – | 0.99 (0.96, 1.00) | – | 0.91 (0.85, 0.95) |
| **18-29 years old** |  |  |  |  |  |  |
| Self-report Vaccinated | 184 | 123 | 0.85 (0.80, 0.90) | – | 0.60 (0.54, 0.65) | – |
| Self-report Unvaccinated | 32 | 568 | – | 0.82 (0.79, 0.85) | – | 0.95 (0.93, 0.96) |
| **30-49 years old** |  |  |  |  |  |  |
| Self-report Vaccinated | 341 | 89 | 0.83 (0.79, 0.86) | – | 0.79 (0.75, 0.83) | – |
| Self-report Unvaccinated | 72 | 559 | – | 0.86 (0.83, 0.89) | – | 0.89 (0.86, 0.91) |
| **> 50 years old** |  |  |  |  |  |  |
| Self-report Vaccinated | 268 | 49 | 0.82 (0.78, 0.86) | – | 0.85 (0.80, 0.88) | – |
| Self-report Unvaccinated | 58 | 291 | – | 0.86 (0.81, 0.89) | – | 0.83 (0.79, 0.87) |
| **C. Region<sup>1</sup>** |  |  |  |  |  |  |
| **Rural** |  |  |  |  |  |  |
| Self-report Vaccinated | 364 | 120 | 0.83 (0.79, 0.86) | – | 0.75 (0.71, 0.79) | – |
| Self-report Unvaccinated | 76 | 781 | – | 0.87 (0.84, 0.89) | – | 0.91 (0.89, 0.93) |
| **Urban** |  |  |  |  |  |  |
| Self-report Vaccinated | 453 | 143 | 0.82 (0.78, 0.85) | – | 0.76 (0.72, 0.79) | – |
| Self-report Unvaccinated | 101 | 993 | – | 0.87 (0.85, 0.89) | – | 0.91 (0.89, 0.92) |

**Abbreviations:** C4: California COVID-19 Case Control study; CAIR: California Immunization Registry; PPV: positive predictive value; NPV: negative predictive value
<sup>1</sup>Regions of participants enrolled in the C4 study were categorized as predominantly rural or predominantly urban according to designations in **Table 1**.

### Interpretation

Among individuals who were unvaccinated at the time of receiving a test for SARS-CoV-2 infection during the period of widespread COVID-19 vaccine availability, we found that COVID-19 vaccination intentions were strongly but imperfectly associated with subsequent initiation of COVID-19 vaccine series. By 5 December 2022, 22% of participants who responded as unsure about receiving COVID-19 vaccines and 13% who expressed unwillingness to receiving COVID-19 vaccines had received at least one dose of COVID-19 vaccine per immunization registry; whereas no record of vaccination was available for 54% of participants who expressed willingness to receive COVID-19 vaccines. Vaccine uptake was fastest among the highest-income households and participants who expressed willingness to receive COVID-19 vaccination. Adjusted hazards of vaccine uptake were higher among school-aged children and teens compared with adults, most notably among the subset of participants who expressed (or had parents/guardians express) hesitancy about receiving COVID-19 vaccination. This finding may reflect the effectiveness of vaccine promotion campaigns, in enhancing COVID-19 vaccine uptake within younger age groups. We identified that a positive SARS-CoV-2 test result predicted lower hazard of COVID-19 vaccination, most strikingly, among individuals who reported being unsure about or unwilling to received vaccine. This suggests that there might be opportunities for outreach to encourage vaccine uptake among individuals who have received a positive COVID-19 test result. Adaptive and dynamic messaging about the strength and durability of infection-induced immunity, and improved efforts to resolve confusion associated with suitable spacing of COVID-19 infection and receipt of COVID-19 vaccination may improve uptake [9].

We did not identify strong evidence of differences in vaccine uptake among unvaccinated individuals according to race/ethnicity, region of residence, anxiety about COVID-19, or opinions about other COVID-19 preventive strategies. No single set of participant-reported reasons for uncertainty or unwillingness to receive COVID-19 vaccine was associated with likelihood of subsequent vaccine uptake. While our findings identify that uncertainty and unwillingness to receive COVID-19 vaccination is not an absolute barrier to subsequent receipt of vaccination, suboptimal vaccine uptake among unvaccinated individuals who expressed willingness to be vaccinated demonstrate gaps in vaccine delivery and/or outreach efforts in California. Associations of vaccine uptake with household income, among participants expressing both uncertainly/unwillingness and willingness to receive COVID-19 vaccination, underscore the need to promote vaccine access and availability in underserved/low-income communities.

Our study complements cross-sectional studies that have characterized vaccine acceptance over time and across communities throughout the pandemic [13–18]. While surveys of vaccine intent can help policymakers understand determinants of vaccine acceptance to better target messaging and resources at populations who may be less willing to initiate vaccination, caution must be used in interpreting these estimates because self-reported acceptance may not translate to vaccine uptake in the real world [19,20]. Indeed, 54% of respondents in our study who expressed willingness to receive COVID-19 vaccination had no evidence of receipt of any vaccine doses within the state-wide immunization rei by late 2021. This observation may indicate social desirability bias among the sample of individuals who consented to participant in a telephone-based questionnaires with public health workers [21]. Our findings are similar to those of a cohort study conducted prior to the widespread availability of COVID-19 vaccines, which likewise identified that 46% of participants who initially expressed enthusiasm about COVID-19 vaccination remained unvaccinated at follow-up during March-April, 2021 [22]. Linking self-reported vaccine hesitancy or willingness with a comprehensive state-wide vaccine registry provided an opportunity to assess alignment of participants’ stated vaccination intentions with real-world vaccine receipt, and to identify predictors of COVID-19 vaccine uptake among participants who initially expressed uncertainty as well as missed opportunities to vaccinate individuals who expressed willingness to receive COVID-19 vaccination.

Few prior studies have assessed the accuracy of self-reported COVID-19 vaccination status. A previous evaluation found agreement between self-reported COVID-19 vaccination status and seropositivity against the SARS-CoV-2 spike protein [22], although such assessments are limited by the fact that seroresponse may also indicate prior infection. Agreement between self-reported vaccination status has been established for other vaccine products, supporting the use of self-reported vaccination status in survey-based research and vaccine effectiveness studies [23]. In our study, specificity of self-reported vaccination status, or the ability to accurately recall not receiving any COVID-19 vaccine doses, was significantly higher than the sensitivity of self-report, or the ability to accurately recall receiving a COVID-19 vaccine dose. Sensitivity of COVID-19 vaccination self-report was notably better among participants referencing a recall aide, especially a COVID-19 vaccination card.

This analysis has several limitations. First, classification of participants with no vaccine record identified in the immunization registry as unvaccinated may be inaccurate; for instance, if individuals received all their vaccine doses outside the state of California. However, this misclassification is likely to be uncommon, given our study was limited to California residents, recommended intervals between receipt of first and second mRNA doses are long, and recommendations for receipt of booster doses were issued during the study period. Second, this study was limited to participants who sought SARS-CoV-2 testing, who may otherwise be more connected to health services and therefore more likely seek vaccination. Third, this analysis evaluated only initiation of the COVID-19 vaccine series which may be an imperfect predictor of willingness to receive subsequent doses needed to maintain or restore immunity to protective levels.

Fourth, this analysis was limited to participants who were unvaccinated throughout the study period and therefore does not estimate determinants of vaccine-uptake across the full population in California; however, predictors of vaccine uptake among the unvaccinated remain important to inform public health policies aimed at improving vaccine coverage. Finally, unmeasured confounding may persist as we were unable to evaluate or control for differences in political affiliation that could further be associated with COVID-19 vaccine initiation [24].

We provide an evaluation of predictors of COVID-19 vaccine uptake and assess the validity of self-reported COVID-19 vaccination status in comparison with a state-wide immunization registry. We identified that self-reported vaccination intent was a strong but imperfect predictor of subsequent vaccine initiation. As no single reason for vaccine hesitancy predicted likelihood of eventual vaccine receipt, public health campaigns addressing multiple factors underlying vaccine hesitancy remain important tools to improve acceptance in hesitant populations.

## Data Availability

Aggregated data may be requested however, subject to approval of the California Health and Human Services Agency - Committee for the Protection of Human Subjects

## Disclaimer

The findings and conclusions in this article are those of the author(s) and do not necessarily represent the views or opinions of the California Department of Public Health or the California Health and Human Services Agency.

## Acknowledgements

We would like to thank all study participants that gave time to complete our survey making possible this work.

## Contributions

KLA – Manuscript writing lead, analysis verification, analysis guidance, conceptualization and JFM – Data matching lead and writing review

NF – Data collection and writing review

LN - Data curation/matching and writing review

RZ – Data curation/matching and writing review

JO – Conceptualization, critical writing edit/review

JPW – Conceptualization, critical writing edit/review

SJ – Conceptualization, funding ascertainment, critical writing edit/review

JAL – Manuscript writing support, analysis guidance, conceptualization and funding ascertainment

JMP – Manuscript writing support, analysis verification, analysis guidance, conceptualization and funding ascertainment

COVID-19 Case-Control Study Team – Data collection and data cleaning

## Notes

### Competing Interest Statement

The authors have declared no competing interest.

### Funding Statement

Work was possible through the support of the Centers of Disease Control and Prevention (CDC) Epidemiology and Laboratory Capacity (ELC) enhancement extension.

### Author Declarations

Study protocol was approved by the California Health and Human Services Agency - Committee for the Protection of Human Subjects project number: 2021-034.

