## Supplementary material for "Real-world uptake of COVID-19 vaccination among individuals expressing vaccine hesitancy: a registry-linkage study": Supplment

^§^	Members of the California COVID-19 Case-Control Study Team include: Helia Samani, Nikolina Walas, Erin Xavier, Diana J. Poindexter, Najla Dabbagh, Michelle M. Spinosa, Shrey Saretha, Adrian F. Cornejo, Hyemin Park, Christine Wan, Miriam I. Bermejo, Amanda Lam, Amandeep Kaur, Ashly Dyke, Diana Felipe, Maya Spencer, Savannah Corredor, Yasmine Abdulrahim, Camilla M. Barbaduomo, Zheng N. Dong, Anna T. Fang, Paulina M. Frost, Timothy Ho, Mahsa H. Javadi, Sophia S. Li, Vivian H. Tran, and Christine Wan

*	KLA and JFM contributed equally to the study.

^†^	JAL and JP jointly supervised the study.

**Figure S1. Kaplan Meier curve**

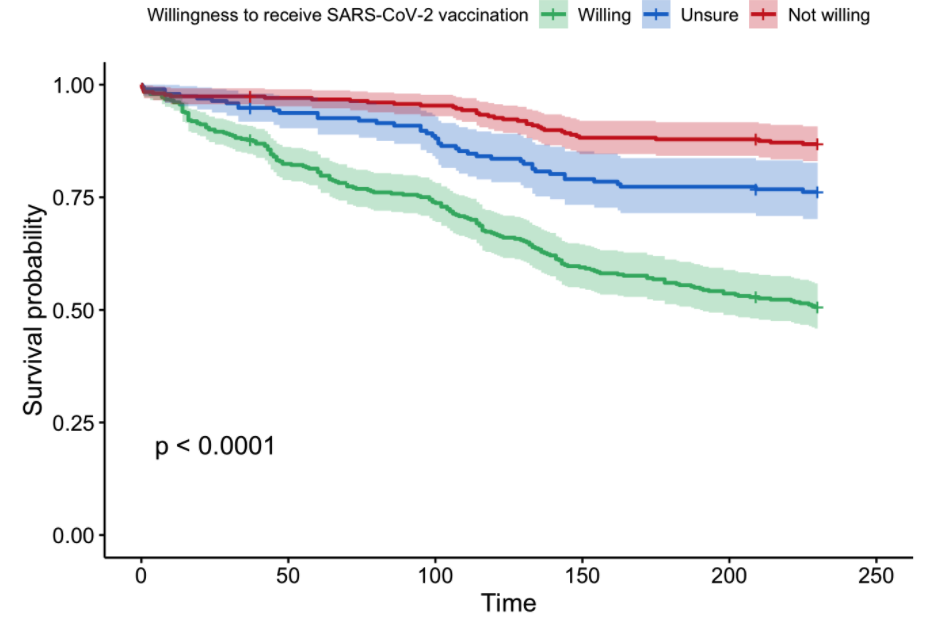

**Table S1: Counties included in each geographic region.**

| County | Region |
| --- | --- |
| Alameda County | San Francisco San Francisco Bay Area |
| Alpine County | Sierras Region |
| Amador County | Sierras Region |
| Butte County | Northern Sacramento Valley |
| Calaveras County | Sierras Region |
| Colusa County | Northern Sacramento Valley |
| Contra Costa County | San Francisco Bay Area |
| Del Norte County | Northwestern California |
| El Dorado County | Sierras Region |
| Fresno County | San Joaquin Valley |
| Glenn County | Northern Sacramento Valley |
| Humboldt County | Northwestern California |
| Imperial County | San Diego and southern border |
| Inyo County | Sierras Region |
| Kern County | San Joaquin Valley |
| Kings County | San Joaquin Valley |
| Lake County | Northwestern California |
| Lassen County | Sierras Region |
| Los Angeles County | Greater Los Angeles area |
| Madera County | San Joaquin Valley |
| Marin County | San Francisco Bay Area |
| Mariposa County | Sierras Region |
| Mendocino County | Northwestern California |
| Merced County | San Joaquin Valley |
| Modoc County | Sierras Region |
| Mono County | Sierras Region |
| Monterey County | Central Coast |
| Napa County | San Francisco Bay Area |
| Nevada County | Sierras Region |
| Orange County | Greater Los Angeles area |
| Placer County | Sierras Region |
| Plumas County | Sierras Region |
| Riverside County | Greater Los Angeles area |
| Sacramento County | Central Valley |
| San Benito County | San Francisco Bay Area |
| San Bernardino County | Greater Los Angeles area |
| San Diego County | San Diego and southern border |
| San Francisco County | San Francisco Bay Area |
| San Joaquin County | San Joaquin Valley |
| San Luis Obispo County | Central Coast |
| San Mateo County | San Francisco Bay Area |
| Santa Barbara County | Central Coast |
| Santa Clara County | San Francisco Bay Area |
| Santa Cruz County | San Francisco Bay Area |
| Shasta County | Northwestern California |
| Sierra County | Sierras Region |
| Siskiyou County | Northwestern California |
| Solano County | San Francisco Bay Area |
| Sonoma County | San Francisco Bay Area |
| Stanislaus County | San Joaquin Valley |
| Sutter County | Northern Sacramento Valley |
| Tehama County | Northern Sacramento Valley |
| Trinity County | Northwestern California |
| Tulare County | San Joaquin Valley |
| Tuolumne County | Sierras Region |
| Ventura County | Greater Los Angeles area |
| Yolo County | Northern Sacramento Valley |
| Yuba County | Northern Sacramento Valley |

**Table S2. Characteristics of records linked between C4 and CAIR.**

|  | Total | Matched^1^ | Unmatched |
| --- | --- | --- | --- |
|  | *N* (%) | *N* (%) | *N* (%) |
|  | *N* = 3031 | *N* = 1080 | *N* = 1951 |
| **Age** |  |  |  |
| 0-6 | 105 (3.5) | 0 (0.0) | 105 (5.4) |
| 7-12 | 135 (4.5) | 6 (0.6) | 129 (6.6) |
| 13-17 | 157 (5.2) | 20 (1.9) | 137 (7.0) |
| 18-29 | 907 (29.9) | 307 (28.4) | 600 (30.8) |
| 30-49 | 1061 (35.0) | 430 (39.8) | 631 (32.3) |
| 50-64 | 462 (15.2) | 210 (19.4) | 252 (12.9) |
| 65+ | 204 (6.7) | 107 (9.9) | 97 (5.0) |
| **Region** |  |  |  |
| *Predominantly Urban Regions* |  |  |  |
| San Francisco Bay Area | 352 (11.6) | 137 (12.7) | 215 (11.0) |
| Greater Los Angeles Area | 330 (10.9) | 110 (10.2) | 220 (11.3) |
| Greater Sacramento Area | 342 (11.3) | 105 (9.7) | 237 (12.1) |
| San Diego and southern border | 338 (11.2) | 110 (10.2) | 228 (11.7) |
| *Predominantly Rural Regions* |  |  |  |
| Central Coast | 362 (11.9) | 151 (14.0) | 211 (10.8) |
| Northern Sacramento Valley | 326 (10.8) | 125 (11.6) | 201 (10.3) |
| San Joaquin Valley | 344 (11.3) | 114 (10.6) | 230 (11.8) |
| Northwestern California | 322 (10.6) | 129 (11.9) | 193 (9.9) |
| Sierras Region | 315 (10.4) | 99 (9.2) | 216 (11.1) |
| **Race/ ethnicity** |  |  |  |
| Non-Hispanic White | 1310 (43.2) | 540 (50.0) | 770 (39.5) |
| Asian | 290 (9.6) | 129 (11.9) | 161 (8.3) |
| Non-Hispanic Black | 135 (4.5) | 30 (2.8) | 105 (5.4) |
| Hispanic (any race) | 834 (27.5) | 256 (23.7) | 578 (29.6) |
| More than 1 race | 308 (10.2) | 84 (7.8) | 224 (11.5) |
| Other race | 72 (2.4) | 22 (2.0) | 50 (2.6) |
| Missing | 82 (2.7) | 19 (1.8) | 63 (3.2) |
| **Use of recall aid** |  |  |  |
| Recall aid not referenced | 304 (10.0) | 130 (12.0) | 174 (8.9) |
| Recall aid referenced | 972 (32.1) | 945 (87.5) | 27 (1.4) |
| Missing recall aid or unvaccinated | 1755 (57.9) | 5 (0.5) | 1750 (89.7) |
| **Month of enrollment (2021)** |  |  |  |
| February | 65 (2.1) | 7 (0.6) | 58 (3.0) |
| March | 550 (18.1) | 106 (9.8) | 444 (22.8) |
| April | 587 (19.4) | 200 (18.5) | 387 (19.8) |
| May | 413 (13.6) | 148 (13.7) | 265 (13.6) |
| June | 295 (9.7) | 99 (9.2) | 196 (10.0) |
| July | 271 (8.9) | 111 (10.3) | 160 (8.2) |
| August | 221 (7.3) | 105 (9.7) | 116 (5.9) |
| September | 167 (5.5) | 79 (7.3) | 88 (4.5) |
| October | 218 (7.2) | 99 (9.2) | 119 (6.1) |
| November | 199 (6.6) | 107 (9.9) | 92 (4.7) |
| December | 45 (1.5) | 19 (1.8) | 26 (1.3) |

**Abbreviations:** C4 = California COVID-19 Case Control study

^1^A participant was considered matched if there was evidence of records of vaccine doses administered after searching for exact matches on zip code of residence, date of birth, and fuzzy matches on first and last name.

**Table S3. Characteristics of participants included in model of vaccine uptake by self-reported willingness to receive vaccination.**

|  | **Willing** | **Unsure** | **Opposed** |
| --- | --- | --- | --- |
|  | *N =* 423 | *N* = 185 | *N* = 278 |
|  | *N* (%) | *N* 9%) | *N* (%) |
| **Age** |  |  |  |
| 5-6 | 20 (4.7) | 5 (2.7) | 5 (1.8) |
| 7-12 | 63 (14.9) | 15 (8.1) | 13 (4.7) |
| 13-17 | 22 (5.2) | 15 (8.1) | 23 (8.3) |
| 18-29 | 142 (33.6) | 65 (35.1) | 82 (29.5) |
| 30-49 | 137 (32.4) | 66 (35.7) | 123 (44.2) |
| 50-64 | 39 (9.2) | 19 (10.3) | 32 (11.5) |
| **Region** |  |  |  |
| *Predominantly Urban Regions* |  |  |  |
| San Francisco Bay Area | 51 (12.1) | 8 (4.3) | 13 (4.7) |
| Greater Los Angeles Area | 49 (11.6) | 21 (11.4) | 36 (12.9) |
| Greater Sacramento Area | 57 (13.5) | 24 (13.0) | 36 (12.9) |
| San Diego and southern border | 65 (15.4) | 23 (12.4) | 28 (10.1) |
| *Predominantly Rural Regions* |  |  |  |
| Central Coast | 46 (10.9) | 14 (7.6) | 25 (9.0) |
| Northern Sacramento Valley | 35 (8.3) | 28 (15.1) | 34 (12.2) |
| San Joaquin Valley | 55 (13.0) | 25 (13.5) | 34 (12.2) |
| Northwestern California | 26 (6.1) | 21 (11.4) | 38 (13.7) |
| Sierras Region | 39 (9.2) | 21 (11.4) | 34 (12.2) |
| **Race/ ethnicity** |  |  |  |
| Non-Hispanic White | 120 (28.4) | 81 (43.8) | 132 (47.5) |
| Asian | 166 (39.2) | 51 (27.6) | 55 (19.8) |
| Non-Hispanic Black | 45 (10.6) | 5 (2.7) | 4 (1.4) |
| Hispanic (any race) | 30 (7.1) | 13 (7.0) | 21 (7.6) |
| More than 1 race | 43 (10.2) | 26 (14.1) | 38 (13.7) |
| Other race | 9 (2.1) | 4 (2.2) | 12 (4.3) |
| Missing | 10 (2.4) | 5 (2.7) | 16 (5.8) |
| **Annual household income** |  |  |  |
| <$50,000 | 134 (31.7) | 48 (25.9) | 74 (26.6) |
| $50,000-$100,000 | 84 (19.9) | 48 (25.9) | 70 (25.2) |
| $100,000-$150,000 | 47 (11.1) | 21 (11.4) | 20 (7.2) |
| >$150,000 | 34 (8.0) | 9 (4.9) | 30 (10.8) |
| Refuse/ missing | 124 (29.3) | 59 (31.9) | 84 (30.2) |
| **Sex** |  |  |  |
| Male | 201 (47.5) | 82 (44.3) | 146 (52.5) |
| Female | 222 (52.5) | 103 (55.7) | 132 (47.5) |
| **Co-morbid conditions** |  |  |  |
| No co-morbidities | 356 (84.2) | 154 (83.2) | 230 (82.7) |
| Co-morbidities | 64 (15.1) | 30 (16.2) | 45 (16.2) |
| Missing | 3 (0.7) | 1 (0.5) | 3 (1.1) |
| **Anxiety about covid** |  |  |  |
| Low anxiety | 255 (60.3) | 135 (73.0) | 229 (82.4) |
| High anxiety | 166 (39.2) | 49 (26.5) | 46 (16.5) |
| Missing | 2 (0.5) | 1 (0.5) | 3 (1.1) |
| **SARS-CoV-2 Test Result** |  |  |  |
| Negative | 137 (32.4) | 64 (34.6) | 94 (33.8) |
| Positive | 286 (67.6) | 121 (65.4) | 184 (66.2) |
| **Agreement with social distancing recommendations** |  |  |  |
| Disagree | 11 (2.6) | 10 (5.4) | 40 (14.4) |
| Neutral | 32 (7.6) | 30 (16.2) | 68 (24.5) |
| Agree | 371 (87.7) | 142 (76.8) | 160 (57.6) |
| Missing | 9 (2.1) | 3 (1.6) | 10 (3.6) |
| **Agreement with face mask policies** |  |  |  |
| Disagree | 11 (2.6) | 25 (13.5) | 56 (20.1) |
| Neutral | 39 (9.2) | 30 (16.2) | 76 (27.3) |
| Agree | 364 (86.1) | 126 (68.1) | 139 (50.0) |
| Missing | 9 (2.1) | 4 (2.2) | 7 (2.5) |

**Table S4. Quantitative bias analysis of vaccine effectiveness estimates.** These estimates were derived from calculations of sensitivity and specificity stratified by SARS-CoV-2 test result status in **Table 5.**

|  | **Cases** | **Controls** | **Odds Ratio** | **Vaccine effectiveness** |
| --- | --- | --- | --- | --- |
|  | *N (%)* | *N* (%) | (95% CI) | % (95 % CI) |
| Self-reported |  |  |  |  |
| Vaccinated | 373 | 707 | 0.37 (0.32, 0.43) | 63 (57-68) |
| Unvaccinated | 1146 | 805 |  |  |
| Bias-corrected |  |  |  |  |
| Vaccinated | 437 | 647 | 0.41 (0.32, 0.51) | 59 (49-68) |
| Unvaccinated | 1168 | 867 |  |  |

**Table S5. Concordance between self-reported first dose product and product listed in CAIR.** Numbers listed in each cell represent the count of individuals for which there was each unique combination of registry (rows) and self-reported (columns) vaccine products.

|  |  | **Self - report (C4 Study)** | | |
| --- | --- | --- | --- | --- |
| **CAIR Registry** |  | Jansen | Moderna | Pfizer |
|  |  | *N* | *N* | *N* |
|  | Jansen | 57 | 0 | 1 |
|  | Moderna | 1 | 311 | 3 |
|  | Pfizer | 2 | 9 | 406 |

**Table S6. Concordance between self-reported dates of first dose and first dose date listed in CAIR2.**

|  | **Total** | **No Vaccine Card or recall aid referenced** | **Vaccine card or recall aid referenced** |
| --- | --- | --- | --- |
|  | *N* = 1456 | *N* = 150 | *N* = 1298 |
| Exact date match | 639 (43.9) | 52 (34.7) | 584 (45.0) |
| ≤14 days | 746 (51.2) | 85 (56.7) | 657(50.6) |
| >14 days | 71 (4.9) | 13 (8.7) | 57 (4.4) |
